# Novel risk models based on screening history results and timing of lung cancer diagnosis: *Post hoc* analysis of the National Lung Cancer Screening Trial

**DOI:** 10.64898/2026.04.12.26350705

**Authors:** Sofia A. Haddan, Asim Waqas, Ghulam Rasool, Matthew B. Schabath

## Abstract

**Background:** Our group previously reported that lung cancer (LC) screening history results and subsequent timing of diagnosis are associated with significant differences in survival outcomes. As a follow-up study, we sought to develop novel personalized risk models that considered screening history for incidence cancers, interval LCs, and prevalence LCs.

**Methods:** Using data from the CT-arm of the NLST, four independent case-control analyses were conducted to develop parsimonious risk models. Controls (n=26,038) were those never diagnosed with LC. The four LC case groups were 270 prevalence LCs, 44 interval LCs, 206 screen-detected LCs (SDLCs) that had a baseline positive screen, and 164 SDLCs that had a baseline negative screen. For each case-control analysis, univariable analyses identified statistically significant covariates from 48 variables and then significant covariates were included into a stepwise backward selection approach to identify a model with the most informative covariates.

**Results:** For prevalence LCs, the model (AUC=0.711) included age, pack-years smoked, BMI, smoking status, smoking onset age, personal history of cancer, family history of LC, alcohol consumption, and milling occupation. For interval LCs, the model (AUC=0.734) included age, smoking status, smoking onset age, cigar smoking, marital status, and asbestos occupation. For baseline positive SDLCs, the model (AUC=0.685) included age, pack-years smoked, BMI, emphysema, chemicals/plastics exposure, and milling occupation. For baseline negative SDLCs, the model (AUC=0.701) included age, pack-years smoked, BMI, smoking status, emphysema, sarcoidosis, and sandblasting occupation.

**Conclusions:** Besides smoking and age, which are inclusion criteria for screening, these models identified other important risk factors which could be used to provide personalized LC risk assessment and screening management.

## INTRODUCTION

The National Lung Screening Trial (NLST) demonstrated a 20% relative reduction in lung cancer mortality for individuals screened by low-dose helical computed tomography (LDCT) compared with chest radiography in a high-risk population of 53,454 current and former smokers aged 55-74 years [1]. While lung cancer screening guidelines in the United States are based on age and smoking history [2-4], these criteria do not take into account other important lung risk factors such as comorbidities, family history, occupational exposure, or other smoking-related factors (e.g., age of initiation). While there are existing risk models [5, 6] that incorporate additional risk factors beyond age and smoking history, and some lung cancer screening guidelines do encourage using such risk calculators [2, 4], these risk models do not consider screening history results, which may provide additional value to personalized risk assessment.

In a prior publication from our group [7], we conducted an in-depth analysis of the CT-arm of the NLST to characterize patient characteristics and survival outcomes of lung cancers diagnosed based on screening history results over the first three screening intervals. A striking finding was that screening history is an independent prognostic factor whereby incidence lung cancers, diagnosed at the follow-up screening intervals, with one or more antecedent negative screens are associated with inferior survival (47.6% 5-year survival) compared to incidence cancers in which only positive screens preceded the lung cancer diagnosis (65.7% 5-year survival). We surmised that the inferior survival associated with incidence lung cancers with one or more antecedent negative screens could be attributed to faster-growing, more biologically aggressive cancers that arose from a lung environment previously lacking a focal abnormality. We also found that the prevalence lung cancers, which are those who have a baseline positive screen that was determined to be cancer, had a 59.9% 5-year survival, and interval cancers, which are diagnosed after a prior negative screening but before the next scheduled screening, were associated with extremely inferior survival (17.3% 5-year survival). As a follow-up to this work, we sought to identify novel personalized risk models for lung cancers based on the screening history results and timing of lung cancer diagnosis, which could be used for personalized management by combining risk factor assessment and screening history.

## MATERIALS AND METHODS

### NLST Study Design, Study Population, and CT Screening Results

This research was approved by the Advarra Institutional Review Board. De-identified NLST CT studies were obtained through the National Cancer Institute Cancer Data Access System[8]. The study design and main findings from NLST have been described previously [1, 9]. Briefly, the NLST was a multicenter randomized clinical trial comparing screening with LDCT versus chest radiography in high-risk individuals. Eligibility criteria included current or former smokers aged 55 to 74 years with a minimum 30 pack-year smoking history. Former smokers must have quit within the past 15 years. Based on the NLST criteria, a positive screening result was defined as having one or more non-calcified nodules or masses with axial diameter measuring ≥ 4 mm. Less commonly, other abnormalities such as adenopathy or pleural effusion can be defined as a positive screening result. Negative screens were defined as CT scans with no abnormalities, minor abnormalities not suspicious for lung cancer, or significant abnormalities not suspicious for lung cancer. Stable abnormalities across all three rounds could be classified as negative screens at the final screen at the discretion of the interpreting radiologist.

Based on a prior publication from our group [7], we restructured the CT arm of the NLST **(Figure 1)** based on screening results at baseline (T0) and the first (T1) and second (T2) follow-up screening intervals. Based on this schema, there were 10 lung cancer case groups that were collapsed into 4 groups for these analyses. Specifically, prevalence lung cancers (green box in **Figure 1)** were diagnosed with lung cancer following a T0 positive screening. Interval lung cancers (yellow boxes in **Figure 1)** were diagnosed after a negative screening at any time between the next scheduled screening interval. A screen-detected lung cancer was defined as an incidence cancer that was identified following a positive screen at the T1 or T2 screening intervals. For this analysis, screen-detected lung cancers were then categorized into two distinct lung cancer case groups according to the baseline screening result (negative or positive). Those with a baseline positive screening who were diagnosed with lung cancer after a follow-up screening interval (T1 or T2) were grouped in one lung cancer case group (blue boxes in **Figure 1)**. Those with a baseline negative screening who were diagnosed with lung cancer after a follow-up screening interval (T1 or T2) were grouped into a separate lung cancer case group (red boxes in **Figure 1)**.

**Figure 1.**
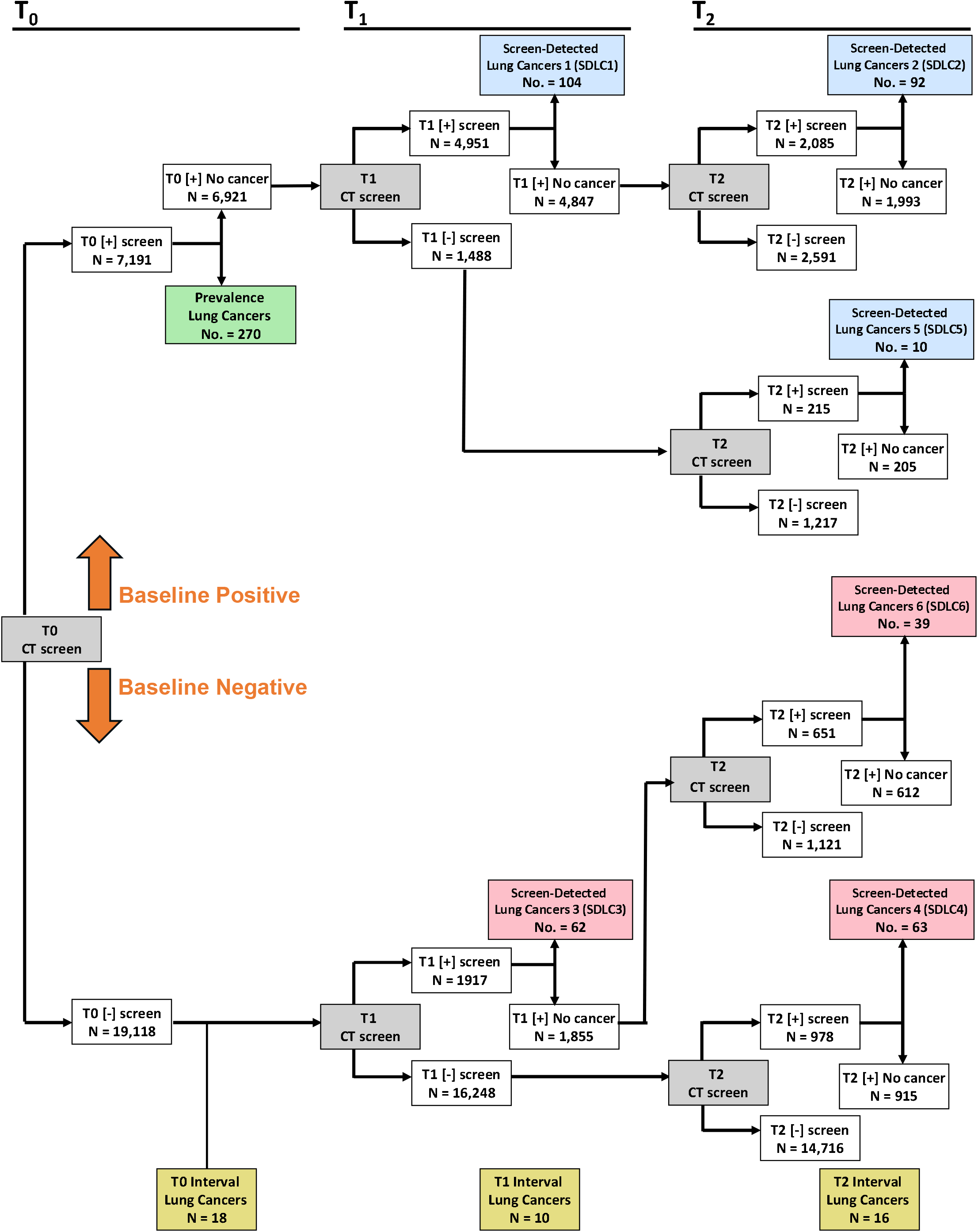
Schema for screening history for all participants spanning three rounds for the LDCT-arm of the NLST.

### Independent variables

***We*** considered 48 independent covariates available from the NLST dataset, including eight demographic variables, six smoking-related variables, two related to alcohol use, fourteen occupational exposure variables, sixteen medical history variables, one variable for family history of lung cancer, and one variable for personal history of any cancer. For this analysis, some of the original NLST variables were re-categorized. Age was categorized into quartiles, and BMI was categorized using the World Health Organization (WHO) guidelines. However, underweight and normal categories were combined due to a low number of participants in the underweight BMI category (e.g., zero participants in this category among interval cancers). The personal history of any cancer was generated by combining the personal history of 15 specific cancer diagnoses. Participants with a history of one or more of these cancers were coded as a “yes,” while a “no” indicated no personal history for any of these cancers. Smoking onset age was categorized into two groups based on the average smoking onset age of 16. For marital status, widowed, separated, and divorced were combined into one category, while never married, married, and not applicable (N/A) were kept as independent categories. The original education was reduced to four categories: (i) high school graduate / GED, 9-11^th^ grade, and 8^th^ grade or less; (ii) associate degree / some college, and post-high school training (excluding college); (iii) graduate school or bachelor’s degree; (iv) other or N/A.

### Statistical Analyses

All statistical analyses were performed using Stata/MP 14.2 for Mac (64-bit Intel). The lung cancer case groups for this analysis were prevalence lung cancers, interval lung cancers, screen-detected lung cancers with a T0 positive screen, and screen-detected lung cancers with a T0 negative screen. The control subjects were participants who were never diagnosed with lung cancer through the original follow-up of the NLST. To assess potential differences in demographic characteristics, Fisher’s exact test was used to test for differences between the control subjects and each of the four lung cancer case groups for categorical variables, and the Student’s t-test was used to test for differences for continuous variables. To identify unique and personalized models associated with lung cancer risk, four independent case-control analyses were performed based on the four different case groups. For each case-control analysis, univariable logistic regression analyses were used to identify statistically significant covariates (p < 0.05) from the 48 covariates. Then, the statistically significant covariates were included into a stepwise backward selection model, using a threshold of ***0*.*1*** for inclusion into the model, which was used to identify the most informative covariates for each case-control analysis. For each final model, we also adjusted for potential confounding, including gender, race, ethnicity, smoke status, and pack-years, where applicable. Area under the curve (AUCs) were generated for each model to measure model performance.

## RESULTS

### Demographics

There were statistically significant differences in the demographic characteristics for all four lung cancer case groups compared to the control subjects for age of enrollment, pack-years smoked, and age of smoking onset **(Table 1)**. Details about the statistically significant differences in the demographic characteristics are presented in the **Supplemental Materials**.

**Table 1.** Demographic and clinical characteristics of lung cancer cases and controls.

|  |  |  |  |  |  |  |
| --- | --- | --- | --- | --- | --- | --- |
|  |  | Lung Cancer Groups |  |  |  |  |
| Characteristic |  | Control<br>Subjects<br>(N = 26,038) | Baseline<br>negative<br>screen-detected lung<br>cancers<br>(N = 164) | Baseline<br>positive<br>screen-detected lung<br>cancers<br>(N = 206) | Prevalence<br>lung cancers<br>(N = 270) | Interval lung<br>cancers<br>(N = 44) |
| Age at enrollment, |  |  |  |  |  |  |
|  | *mean (SD)* | 61.4 (5.0) | 63.4 (5.1)*** | 63.7 (5.1)*** | 63.8 (5.5)*** | 63.0 (5.1)** |
|  | *categorical, N (%)* |  |  |  |  |  |
|  | ≤ 56 | 5,195 (20.0) | 14 (8.5)*** | 17 (8.3)*** | 30 (11.1)*** | 5 (11.4) |
|  | 57-59 | 6,064 (23.3) | 31 (18.9) | 33 (16.0) | 45 (16.7) | 8 (18.2) |
|  | 60-64 | 7,961 (30.6) | 55 (33.5) | 64 (31.1) | 76 (28.2) | 14 (31.8) |
|  | ≥ 65 | 6,818 (26.2) | 64 (39.0) | 92 (44.7) | 119 (44.1) | 17 (38.6) |
| Gender, N (%) |  |  |  |  |  |  |
|  | Female | 10,682 (41.0) | 58 (35.4) | 89 (43.2) | 113 (41.9) | 11 (25.0)* |
|  | Male | 15,356 (59.0) | 106 (64.6) | 117 (56.8) | 157 (58.1) | 33 (75.0) |
| Race, N (%) |  |  |  |  |  |  |
|  | White | 23,657 (90.9) | 147 (89.6) | 195 (94.7) | 250 (92.6) | 40 (90.9) |
|  | Non-white, NA, or<br>more than one race | 2,381 (9.1) | 17 (10.4) | 11 (5.34) | 20 (7.4) | 4 (9.1) |
| Ethnicity, N (%) |  |  |  |  |  |  |
|  | Neither Hispanic nor<br>Latino | 25,408 (97.6) | 163 (99.4) | 205 (99.5) | 260 (96.3) | 43 (97.7) |
|  | Hispanic or Latino | 472 (1.8) | 1 (0.6) | 0 (0) | 6 (2.2) | 0 (0) |
|  | Missing | 158 (0.6) | 0 (0) | 1 (0.5) | 4 (1.5) | 1 (2.3) |
| Marital status, N (%) |  |  |  |  |  |  |
|  | Married | 17,371 (66.7) | 112 (68.3) | 131 (63.6) | 178 (65.9) | 23 (52.3)*** |
|  | Never Married | 1,229 (4.7) | 4 (2.4) | 9 (4.4) | 13 (4.8) | 0 (0) |
|  | Widowed/<br>separated/ divorced | 7,305 (28.1) | 48 (29.3) | 65 (31.6) | 79 (29.3) | 19 (43.2) |
|  | Unknown | 133 (0.5) | 0 (0) | 1 (0.5) | 0 (0) | 2 (4.6) |
| Family history of lung cancer, N (%) |  |  |  |  |  |  |
|  | No | 20,397 (78.3) | 126 (76.8) | 156 (75.7) | 191 (70.7)*** | 37 (84.1) |
|  | Yes | 5,641 (21.7) | 38 (23.2) | 50 (24.3) | 79 (29.3) | 7 (15.9) |
| Personal history of any cancer, N (%) |  |  |  |  |  |  |
|  | No | 24,921 (96.0) | 155 (94.5) | 193 (93.7) | 245 (90.7)*** | 42 (97.7) |
|  | Yes | 1,048 (4.0) | 9 (5.5) | 13 (6.3) | 25 (9.3) | 1 (2.3) |
| Smoking status, N (%) |  |  |  |  |  |  |
|  | Former | 13,566 (52.1) | 63 (38.4)*** | 97 (47.1) | 121 (44.8)* | 15 (34.1)* |
|  | Current | 12,472 (47.9) | 101 (61.6) | 109 (52.9) | 149 (55.2) | 29 (65.9) |
| Pack-years smoked, mean (SD) |  |  |  |  |  |  |
|  | Overall | 55.8 (23.8) | 67.8 (27.9)*** | 64.3 (26.8)*** | 65.5 (29.8)*** | 68.0 (27.7)*** |
|  | Former smokers | 56.3 (25.0) | 67.5 (29.2) | 65.7 (27.4) | 65.6 (30.5) | 69.4 (31.3) |
|  | Current smokers | 55.2 (22.5) | 68.0 (27.2) | 63.1 (26.4) | 65.5 (29.3) | 67.3 (26.2) |

Table 1. Continued
| Characteristic |  | Control Subjects<br>(N = 26,038) | Lung Cancer Groups |  |  |  |
| --- | --- | --- | --- | --- | --- | --- |
|  |  |  | Baseline negative screen-detected lung cancers<br>(N = 164) | Baseline positive screen-detected lung cancers<br>(N = 206) | Prevalence lung cancers<br>(N = 270) | Interval lung cancers<br>(N = 44) |
| **Pack-years smoked, categorical, N (%)** |  |  |  |  |  |  |
|  | <=38 | 5,952 (22.9) | 18 (11.0)*** | 22 (10.7)*** | 19 (7.0)*** | 2 (4.6)** |
|  | 39-47 | 6,808 (26.2) | 31 (18.9) | 39 (18.9) | 69 (25.6) | 13 (29.6) |
|  | 48-65 | 6,464 (24.8) | 37 (22.6) | 63 (30.6) | 80 (29.6) | 10 (22.7) |
|  | >=66 | 6,814 (26.2) | 78 (47.6) | 82 (39.8) | 102 (37.8) | 19 (43.2) |
| **Age initiation of smoking, mean (SD)** |  |  |  |  |  |  |
|  | Overall | 16.7 (3.7) | 16.1 (3.8)* | 16.2 (3.1)* | 16.2 (3.7)* | 15.0 (3.7)** |
|  | Former smokers | 16.6 (3.5) | 16.6 (4.1) | 16.3 (2.7) | 16.1 (3.7) | 15.8 (2.6) |
|  | Current smokers | 16.9 (3.9) | 15.8 (3.6) | 16.1 (3.5) | 16.3 (3.8) | 14.7 (3.7) |
| **BMI, mean (SD)** |  |  |  |  |  |  |
|  |  | 27.9 (5.0) | 26.6 (4.6)*** | 26.9 (5.1)** | 26.8 (4.4)*** | 27.1 (4.3) |
| **BMI, categorical, N (%)** |  |  |  |  |  |  |
|  | Underweight / Healthy Weight (<=24) | 7500 (28.8) | 53 (32.3)** | 75 (36.4)** | 104 (38.5)*** | 15 (34.1) |
|  | Overweight (25-29) | 10,973 (42.1) | 83 (50.6) | 86 (41.8) | 111 (41.1) | 15 (34.1) |
|  | Obesity (>=30) | 7,565 (29.1) | 28 (17.1) | 45 (21.8) | 55 (20.4) | 14 (31.8) |

\* P < 0.05; \*\* P < 0.01; \*\*\* P < 0.001
**Original data for BMI before collapsing underweight + healthy weight categories:**
Interval lung cancer cases:
-Underweight: 0
Healthy weight: 15
Overweight: 15
Obese: 14

Figure 2 graphically represents the covariates for each of the risk models from the four case-control analyses.

**Figure 2.**
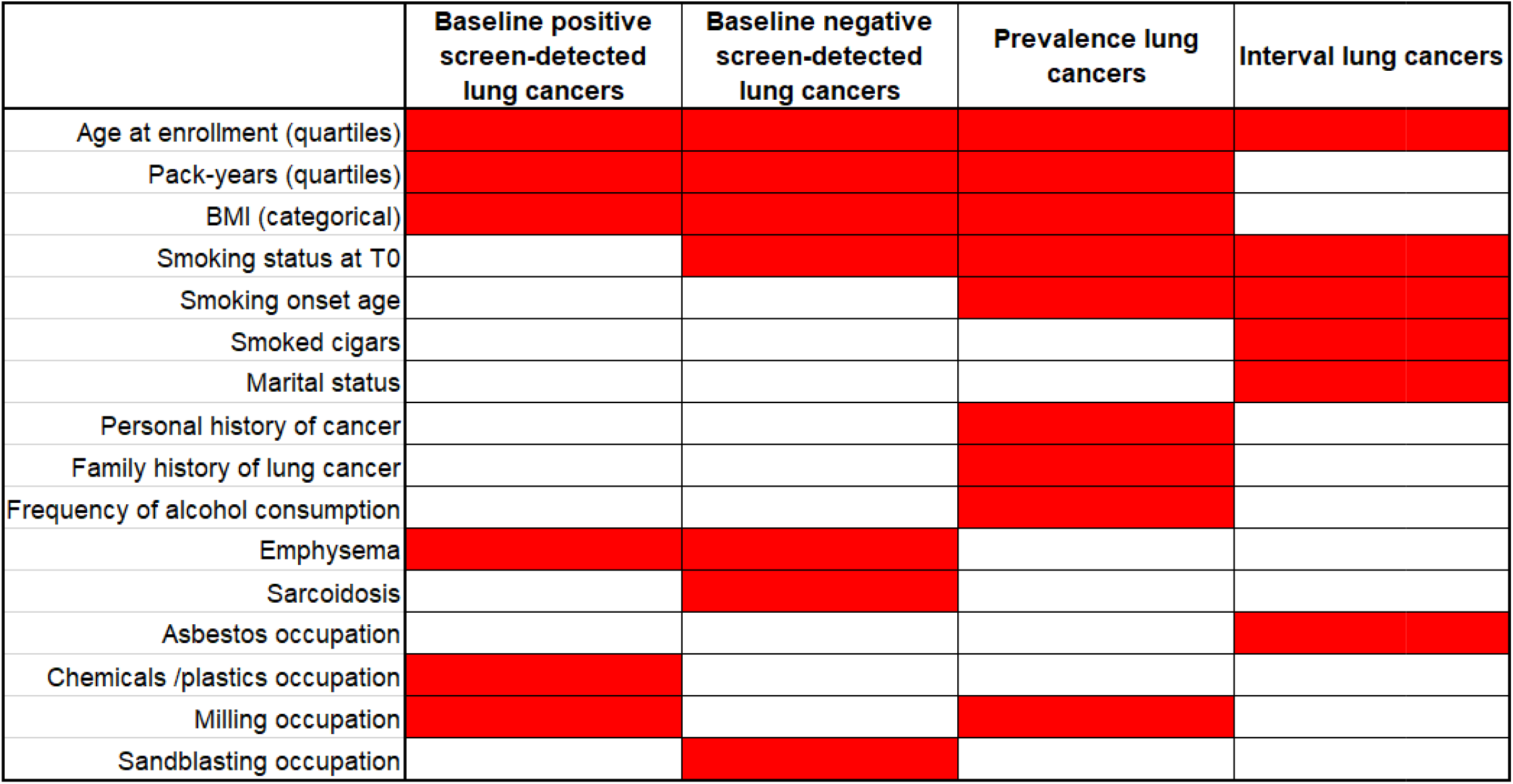
Graphical representation of the risk models from the four case-control analyses.

### Incidence Lung Cancers Diagnosed Following a Baseline Positive Screen

Univariable analyses revealed the following variables to be statistically significantly associated with lung cancer risk for incidence lung cancers diagnosed following a baseline positive screen **(Supplemental Table 1):** age at enrollment, pack-years smoked, BMI, past medical history of COPD, past medical history of emphysema, occupational exposure to chemicals/plastics, occupational exposure to flour feed/grain milling, and occupational exposure to welding. After inclusion of the statistically significant univariable covariates into the stepwise backward elimination model, the final risk model **(Table 2)** included age at enrollment, pack-years smoked, BMI, past medical history of emphysema, occupational exposure to chemicals/plastics, and occupational exposure to flour feed/grain milling. Specifically, compared to the first age quartile (≤ 56), individuals in the second quartile of age (57-59) exhibited a 59% (OR = 1.59; 95% CI: 0.88 – 2.87) increased risk of lung cancer, individuals in the third quartile (60-64) had a 2.21-fold (OR = 2.21; 95% CI: 1.29 – 3.79) increased risk of lung cancer, and individuals in the fourth quartile (≥ 65) had 3.35-fold (OR = 3.35; 95% CI: 1.98 – 5.66) increased risk of lung cancer. For pack-years smoked, compared to the first quartile (≤ 38 pack-years) the second quartile (39-47 pack years smoked) exhibited a 37% (OR 1.37; 95% CI: 0.81 – 2.31) increased risk of lung cancer, the third quartile (48-65 pack years smoked) revealed a 2.17-fold (95% CI: 1.33 – 3.55) increased risk of lung cancer, and fourth quartile (≥ 66 pack years smoked) was associated with a 2.59-fold (95% CI: 1.60 – 4.19) increased risk of lung cancer. For BMI, the underweight/healthy was set as the reference. Overweight BMI (25 – <30) was associated with a borderline significant 23% (95% CI: 0.56 – 1.05) decreased risk of lung cancer, while obesity (≥30) was associated with a 41% (95% CI: 0.40 – 0.85) decreased risk of lung cancer. Past medical history of emphysema was associated with a 52% (OR = 1.52; 95% CI: 1.02 – 2.26) increased risk of lung cancer, occupational exposure to chemicals was associated with 73% (OR = 1.73; 95% CI: 1.11 – 2.72) increased risk of lung cancer, and occupational exposure to flour feed grain milling was associated with 2.30- fold (95% CI: 1.01 – 5.28) increased risk of lung cancer.

**Table 2.**
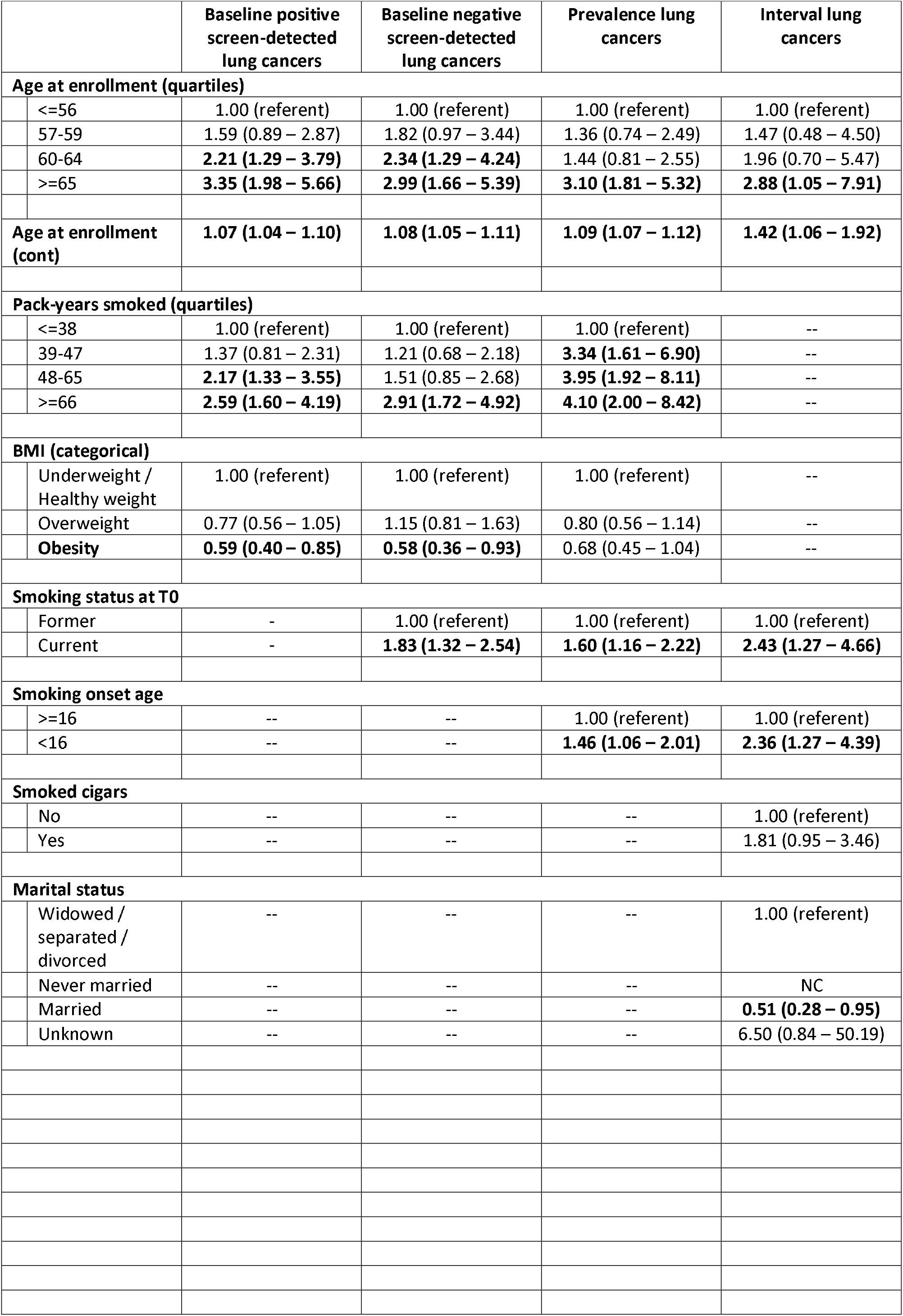
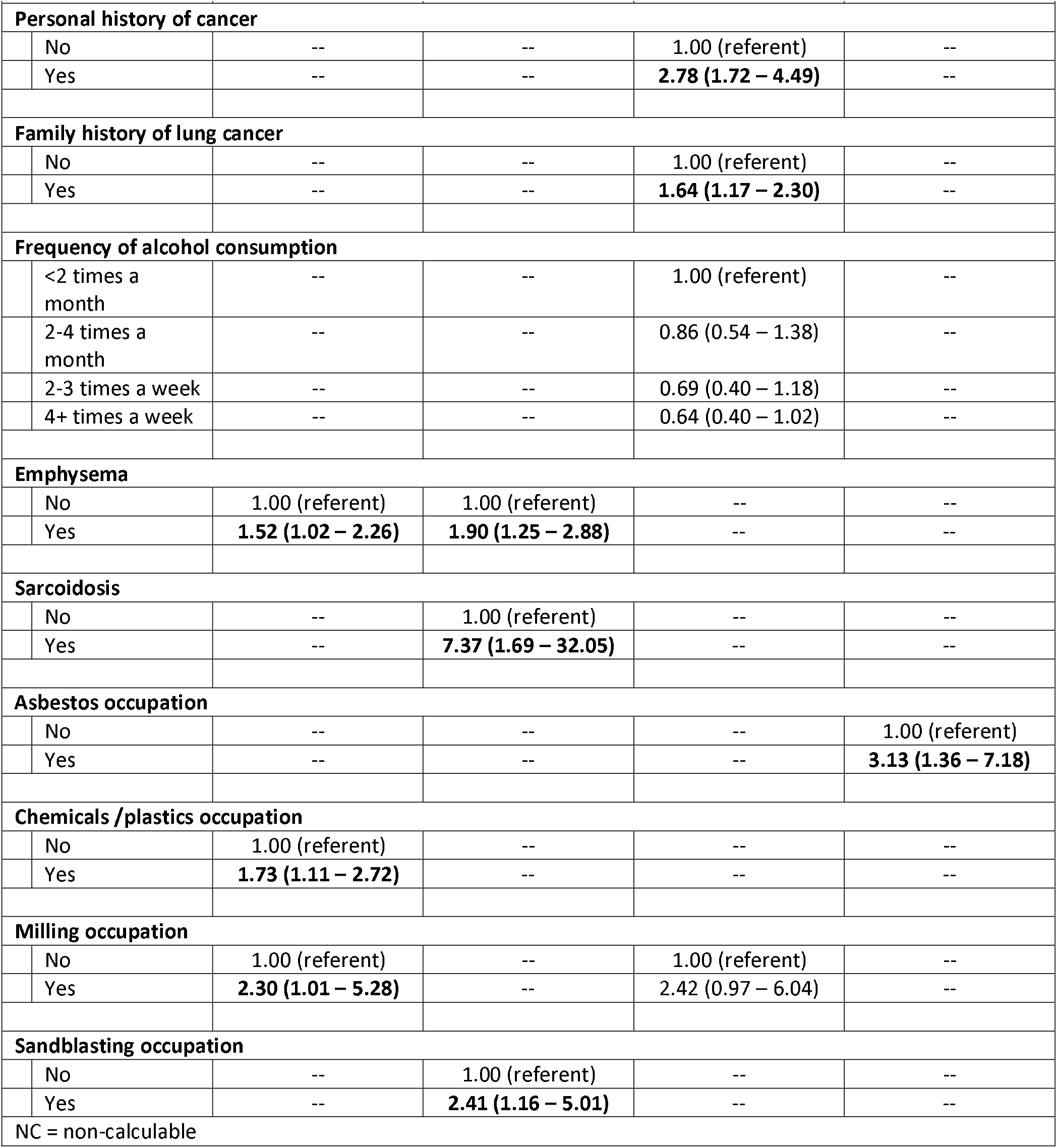
Lung cancer risk models based on screening history.

### Incidence Lung Cancers Diagnosed Following a Baseline Negative Screen

Univariable analyses revealed the following variables to be statistically significantly associated with lung cancer risk for incidence lung cancers diagnosed following a baseline negative screen **(Supplemental Table 1):** age at enrollment, pack-years smoked, BMI, smoking status at T0, smoking onset age, past medical history of COPD, past medical history of emphysema, past medical history of sarcoidosis, and occupation exposure to sandblasting. After inclusion of the statistically significant univariable covariates into a stepwise backward elimination model, the final risk model **(Table 2)** included age at enrollment, pack-years smoked, BMI, smoking status at T0, past medical history of emphysema, past medical history of sarcoidosis, and occupational exposure to sandblasting. Specifically, compared to the first age quartile (≤56 years), individuals in the second quartile of age (57 – 59) exhibited an 82% (95% CI: 0.97 – 3.44) increased risk of lung cancer, individuals in the third quartile of age (60 – 64) exhibited a 2.34-fold (95% CI: 1.29 – 4.24) increased risk of lung cancer, and individuals in the fourth quartile of age (≥65) exhibited a 2.99-fold (95% CI: 1.66 – 5.39) increased risk of lung cancer. For pack-years smoked, compared to the first quartile (≤38 pack-years), individuals in the second quartile (39-47 pack-years) exhibited a 21% (95% CI: 0.68 – 2.18) increased risk of lung cancer, individuals in the third quartile (48-65 pack years) exhibited a 51% (95% CI: 0.85 – 2.68) increased risk of lung cancer, and individuals in the fourth quartile (≥66 pack-years) exhibited a 2.91-fold (95% CI: 1.72 – 4.92) increased risk of lung cancer. For BMI, compared to the reference category (25 - <30), individuals in the overweight category exhibited a 15% (95% CI: 0.81 – 1.63) increased risk of lung cancer, and individuals in the obesity category (BMI: ≥30) saw a 42% (95% CI: 0.36 – 0.93) decreased risk of lung cancer. For smoking status at T0, compared to former smokers, individuals who were current smokers saw an 83% (95% CI: 1.32 – 2.54) increased risk of lung cancer. Individuals with a past medical history of emphysema exhibited a 90% (95% CI: 1.25 – 2.88) increased risk of lung cancer. Individuals with a past medical history of sarcoidosis exhibited a 7.37-fold (95% CI: 1.69 – 32.05) increased risk of lung cancer. Individuals with a current or history of occupational exposure to sandblasting exhibited a 2.41-fold (95% CI: 1.16 – 5.01) increased risk of lung cancer.

### Prevalence Lung Cancers

Univariable analyses revealed the following variables to be statistically significantly associated with lung cancer risk for prevalence lung cancers **(Supplemental Table 1):** age at enrollment, education, pack-years smoked, BMI, personal history of any cancer, family history of lung cancer, smoking status at T0, smoking onset age, past medical history of COPD, and past medical history of emphysema. After inclusion of the statistically significant univariable covariates into a stepwise backward elimination model, the final risk model **(Table 2)** included age at enrollment, pack-years smoked, BMI, smoking status at T0, smoking onset age, personal history of cancer, family history of lung cancer, frequency of alcohol consumption, and occupational exposure to flour feed / grain milling.

Specifically, compared to the first age quartile (≤56 years), individuals in the second quartile of age (57 – 59) exhibited a 36% (95% CI: 0.74 – 2.49 increased risk of lung cancer, individuals in the third quartile of age (60 – 64) exhibited a 44% (95% CI: 0.81 – 2.55) increased risk of lung cancer, and individuals in the fourth quartile (ages ≥65) exhibited a 3.10-fold (95% CI: 1.81 – 5.32) increased risk of lung cancer. For pack-years smoked, compared to the first quartile (≤38 pack-years), individuals in the second quartile (39-47 pack-years) exhibited a 3.34-fold (95% CI: 1.61 – 6.90) increased risk of lung cancer, individuals in the third quartile (48-65 pack years smoked) exhibited a 3.95-fold (95% CI: 1.92 – 8.11) increased risk of lung cancer, and individuals in the fourth quartile (≥66 pack years smoked) exhibited a 4.10-fold (95% CI: 2.00 – 8.42) increased risk of lung cancer. For BMI, compared to the reference category (25 - <30), individuals in the overweight category exhibited a 20% (95% CI: 0.56 – 1.14) decreased risk of lung cancer, and individuals in the obesity category (BMI: ≥30) exhibited a 32% (95% CI: 0.45 – 1.04) decreased risk of lung cancer. For smoking status at T0, compared to former smokers, current smokers exhibited a 60% (95% CI: 1.16 – 2.22) increased risk of lung cancer. For smoking onset age, compared to individuals who began smoking after the age of 16, individuals who started smoking before the age of 16 exhibited a 46% (95% CI: 1.06 – 2.01) increased risk of lung cancer. Individuals with a personal history of any cancer exhibited a 2.78-fold (95% CI: 1.72 – 4.49) increased risk of lung cancer. Individuals with a family history of lung cancer exhibited a 64% (95% CI: 1.17 – 2.30) increased risk of lung cancer. For frequency of alcohol consumption, compared to individuals who consumed alcohol less than two times a month, individuals who consumed alcohol from two to four times a month exhibited a 14% (95% CI: 0.54 – 1.38) decreased risk of lung cancer, individuals who consumed alcohol 2-3 times a week exhibited a 31% (95% CI: 0.40 – 1.18) decreased risk of lung cancer, and individuals who consumed alcohol four or more times per week exhibited a 36% (95% CI: 0.40 – 1.02) decreased risk of lung cancer. Occupational exposure to flour feed / grain milling was associated with a 2.42-fold (95% CI: 0.97 – 6.04) increased risk of lung cancer.

### Interval Lung Cancers

Univariable analyses revealed the following variables to be statistically significantly associated with lung cancer risk for interval lung cancers **(Supplemental Table 1):** age at enrollment, gender, pack-years smoked, marital status, history of smoking cigars, smoking status at T0, smoking onset age, past medical history of asbestosis, occupational exposure to asbestos, and occupational exposure to sandblasting. After inclusion of the statistically significant univariable covariates into a stepwise backward elimination model, the final risk model **(Table 2)** included age at enrollment, smoking status at T0, smoking onset age, history of smoking cigars, marital status, and occupational exposure to asbestos. Specifically, compared to the first age quartile (<=56 years), individuals in the second quartile of age (57 – 59) exhibited a 47% (95% CI: 0.48 – 4.50) increased risk of lung cancer, individuals in the third quartile of age (60 – 64) exhibited a 96% (95% CI: 0.70 – 5.47) increased risk of lung cancer, and individuals in the fourth quartile (ages ≥65) exhibited a 2.88-fold (95% CI: 1.05 – 7.91) increased risk of lung cancer. For smoking status at T0, compared to former smokers, current smokers exhibited a 2.43-fold (95% CI: 1.27 – 4.66) increased risk of lung cancer. For smoking onset age, compared to individuals who began smoking after the age of 16, individuals who began smoking before the age of 16 exhibited a 2.36-fold (95% CI: 1.27 – 4.39) increased risk of lung cancer. Individuals with a personal history of any cancer exhibited a 2.78-fold (95% CI: 1.72 – 4.49) increased risk of lung cancer. Individuals with a history of smoking cigars exhibited an 81% (95% CI: 0.95 – 3.46) increased risk of lung cancer. For marital status, compared to those who were widowed, separated, or divorced, individuals who were married exhibited a 49% (95% CI: 0.28 – 0.95) decreased risk of lung cancer. Occupational exposure to asbestos was associated with a 3.13-fold (95% CI: 1.36 – 7.18) increased risk of lung cancer.

### Summary of risk models

Figure 2 graphically represents the covariates for each of the risk models from the four case-control analyses.

### Adjusted multivariable risk models

Supplemental Table 2 presents the final risk models adjusted for potential confounders including, where relevant, gender, race, ethnicity, smoke status, and pack-year smoked. The adjusted models were largely similar to the unadjusted lung cancer risk models presented in Table 2.

### Survival Analysis

In our prior publication [7], we did not analyze overall survival for case groups SDLC5 (blue box in Figure 1) and SDLC6 (red box in Figure 1). Consistent with the prior findings, incidentally detected lung cancers with one or more preceding negative screens (red line in Figure 3) are associated with poorer survival outcomes compared to those preceded by only positive screens (blue line in Figure 3).

**Figure 3.**
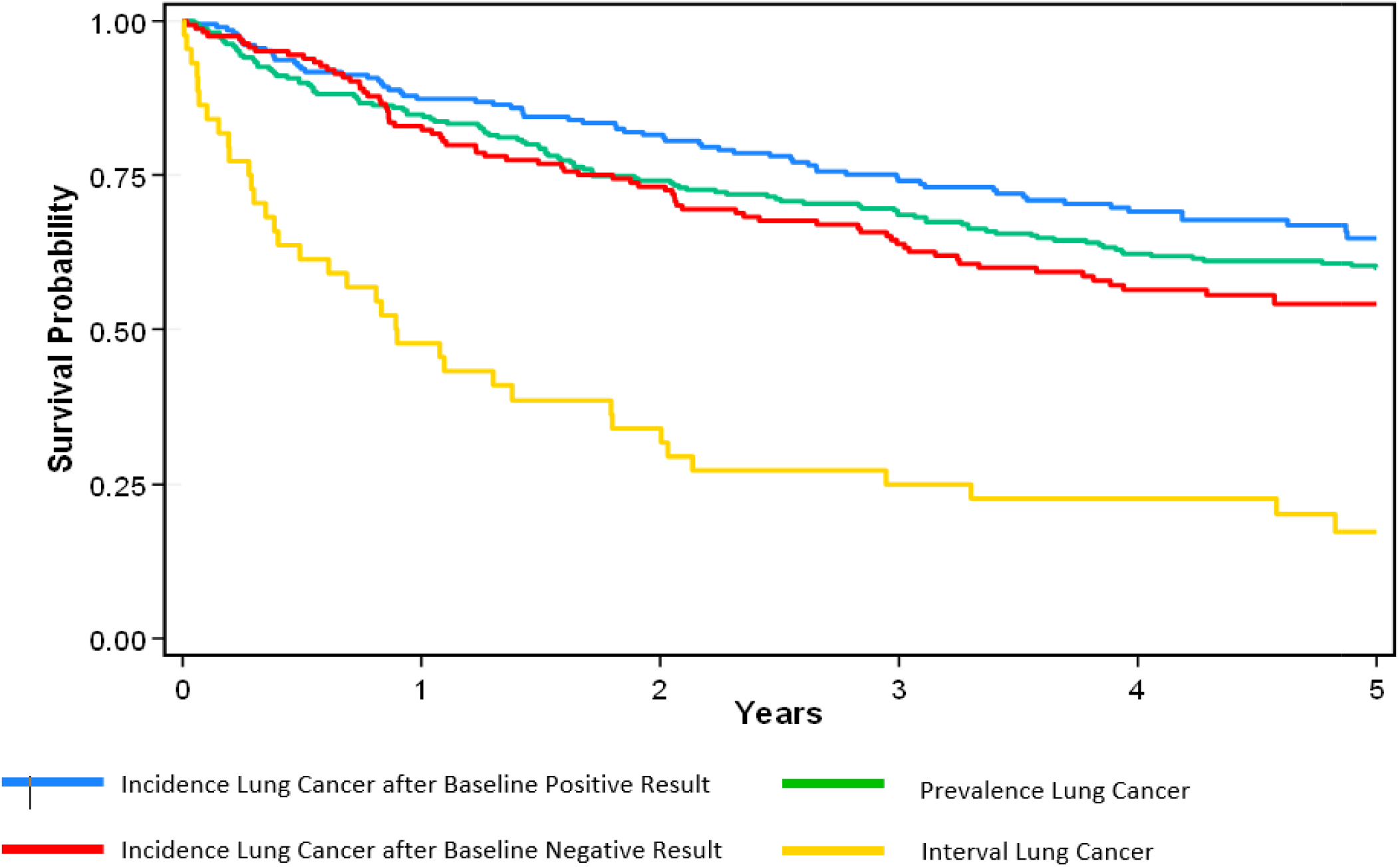
Kaplan-Meier estimates for overall survival in the prevalence-, interval-, and screen-detected (incidence) lung cancers.

## DISCUSSION

In this ***post hoc*** analysis on the NLST, we sought to identify personalized risk models based in lung cancer screening that considered screening history, timing of lung cancer diagnosis, and person-level data beyond age and smoking history. We deployed a rigorous epidemiologic model-building approach to identify the most informative covariates from all available demographic and risk factor data in the NLST to generate four parsimonious lung cancer risk models. The models incorporate additional important risk factors for lung cancer, including demographics, other smoking covariates, occupational exposures, medical history, alcohol use, personal history of cancer, and family history of lung cancer. The models could be used to provide personalized risk assessment and screening management by identifying high-risk individuals using information easily obtained in the lung cancer screening setting.

Age was the only covariate found to be informative in all four models. Old age was consistently associated with an increased risk of lung cancer. Other than smoking history, age is one of the most important and well-established risk factors present in nearly all lung cancer risk models [10] including, but not limited to, Bach model [11], Liverpool Lung Project (LLP) model [12], PLCOm2012 model [5], Two-Stage Clonal Expansion (TSCE) model[13], and the Pittsburgh Predictor[14]. Pack-years smoked and smoking status were found to be informative in three of the four models. While pack-years smoked was not found in the model for interval lung cancers, smoking status and cigar smoked were included. Because interval cancers were the smallest case group, there may not have been the statistical power to detect statistically significant odds ratios for the pack-years variable, which was analyzed by quartiles. While smoking status was not found in the model for baseline/prevalence lung cancer, pack-years was included and emphysema which is strongly linked to cigarette smoking was also included [15].

BMI was found to be informative in three of the models, revealing an inverse association between high BMI and lung cancer risk. Similar to pack-years smoked, BMI was not included in the interval cancers model, which is likely attributed to the small sample size and statistical power to detect statistically significant odds ratios. While BMI is included in the PLCOm2012 risk model [5], it is not present in other models [5, 12-14].

Consistent with the PLCOm2012 model [5] and our findings, prior meta- and pooled-analyses [16,17] revealed that leanness is associated with an increased risk of lung cancer. Specifically, a pooled analysis of 10 prospective cohort studies in Japan revealed that leanness (BMI<18.5) was associated with a 35% increased risk of lung cancer (95% CI: 1.16 – 1.57), and overweight and obesity were associated with a lower risk of lung cancer, with hazard ratios (HR) of 0.77 (95% CI: 0.71 – 0.84) and 0.69 (95% CI: 0.45 – 1.07), respectively [16]. A pooled analysis with cohorts based in the United States, Europe, China, and Singapore showed a decreased risk of lung cancer for those who were overweight (OR = 0.77, 95% CI: 0.68 – 0.86) and obese (OR=0.69, 95% CI: 0.59 - 0.82) [17]. This inverse association could be attributed to smoking, because nicotine, the major addictive component of cigarette smoking, is associated with reduced appetite and alters eating patterns, typically resulting in reduced body weight [18, 19]. However, when we adjusted for smoking history, the associations with BMI were generally consistent. As such, the observed inverse associations with BMI could be causal or attributed to unmeasured confounding.

Age of initiation of smoking, self-reported history of emphysema, and milling occupation were all found to be relevant in two of the four models, and there were seven covariates that were found only in one model. Given that these four models have more unique risk factors rather than shared risk factors **(Figure 2)**, it suggests that a single risk model may not provide personalized follow-up and screening management in lung cancer screening. Certainly, prior models [5, 10-14] were not developed considering screening history and timing of the lung cancer diagnosis, which add to the novelty of the analyses conducted in this study. Additionally, many of the other studies did not have, or did not consider, the sizeable and diverse number of risk factors that are available in the NLST. For example, simpler risk models, such as the Pittsburgh Predictor uses only four risk factors [14], of which three are smoking-related: duration of smoking, smoking status, and smoking. Personal history of any cancer and family history of lung cancer, found only in the prevalence model, are included in the Liverpool Lung Project model and the PLCOm2012 model.

We do acknowledge some limitations of this study. The generalizability of our results to screening populations outside the NLST eligibility criteria is unknown. Despite conducting multivariable analyses, we cannot account for biases from unknown confounders and unmeasured covariates. Self-report data, which was used for the model building, is subject to recall biases and misclassification. The screening results to differentiate positive vs. negative findings were based on the original NLST guidelines, which differ from current guidelines in the United States [2]. Despite these limitations, the results from this analysis emphasize the importance of considering screening history in the development of personalized risk models.

This is one of the first studies to develop personalized risk models that take into account screening history and putative and causal lung cancer risk factors. By doing so, this study identified personalized lung cancer risk models that integrate critical risk factors beyond age and smoking history, including other smoking covariates, demographic information, alcohol use, medical history, family history of lung cancer, and occupational exposures. These models offer a nuanced approach to identifying high-risk individuals for lung cancer and provide a basis for personalized screening strategies and preventative measures. By personalizing the lung cancer screening and management process for individuals, healthcare providers can enhance early detection and implement timely intervention strategies. Integrating these models in a clinical setting has the potential to improve patient outcomes and make a meaningful contribution to lung care prevention and care.

## Data Availability

None of the authors in the manuscript are affiliated with the NCI and none of the authors own the dataset. All de-identified NLST data files are available from the National Cancer Institute (NCI) Cancer Data Access System (CDAS). We did not generate any proprietary variables. As such, the same data used in our analyses are available to all researchers by submitting an application at CDAS.

## Supplemental Materials

### Results

Demographic and clinical characteristics of lung cancer cases and controls are presented **(Table 1)**. Specifically, controls (mean = 61.4 [SD 5.0]) were statistically significantly younger compared to baseline negative screen-detected lung cancers (mean = 63.4 [SD 5.1]), baseline positive screen-detected lung cancers (mean = 63.7 [SD 5.1]), prevalence lung cancers (mean = 63.8 [SD 5.5]), and interval lung cancers (mean = 63.0 [5.1]). Controls (mean = 55.8 [SD 23.8]) reported a statistically significantly lower number of pack-years smoked compared to baseline negative screen-detected lung cancers (mean = 67.8 [SD 27.9]), baseline positive screen-detected lung cancers (mean = 64.3 [SD 26.8]), prevalence lung cancers (mean = 65.5 [SD 29.8]), and interval lung cancers (mean = 68.0 [27.7]). Controls (mean = 16.7 [SD 3.7]) also reported a statistically significantly higher mean age of smoking onset compared to baseline negative screen-detected lung cancers (mean = 16.1 [SD 3.8]), baseline positive screen-detected lung cancers (mean = 16.2 [SD 3.1]), prevalence lung cancers (mean = 16.2 [SD 3.7]), and interval lung cancers (mean = 15.0 [SD 3.7]). For smoking status, control subjects (47.9%) had a statistically significantly lower percentage of current smokers compared to baseline negative screen-detected lung cancers (61.6%, p-value =0.001), prevalence lung cancers (55.2%, p-value =0.020), and interval lung cancers (65.9%, p-value =0.022). With respect to BMI, controls (mean = 27.9 [SD 5.0]; p-value = 0.002) had significantly higher BMIs on average compared to baseline negative screen-detected lung cancers (mean = 26.6 [SD 4.6]; p-value =0.002), baseline positive screen-detected lung cancers (mean = 26.9 [SD 5.1]; p-value = 0.021), and prevalence lung cancers (mean = 26.8 [SD 4.4]; p-value <0.001). There were no statistically significant differences between controls and the four lung cancer case groups for race and ethnicity.

**Supplemental Table 1.**
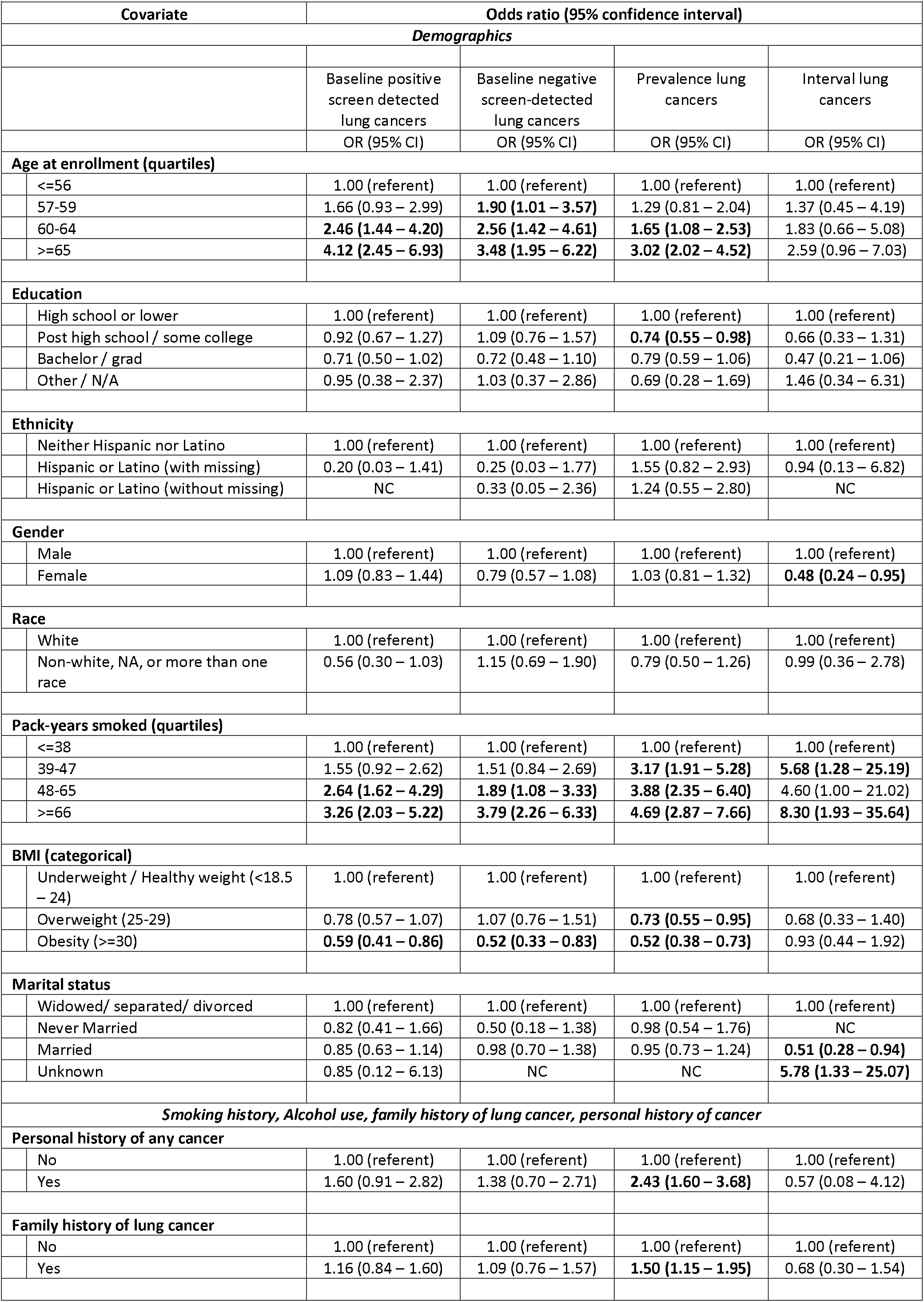
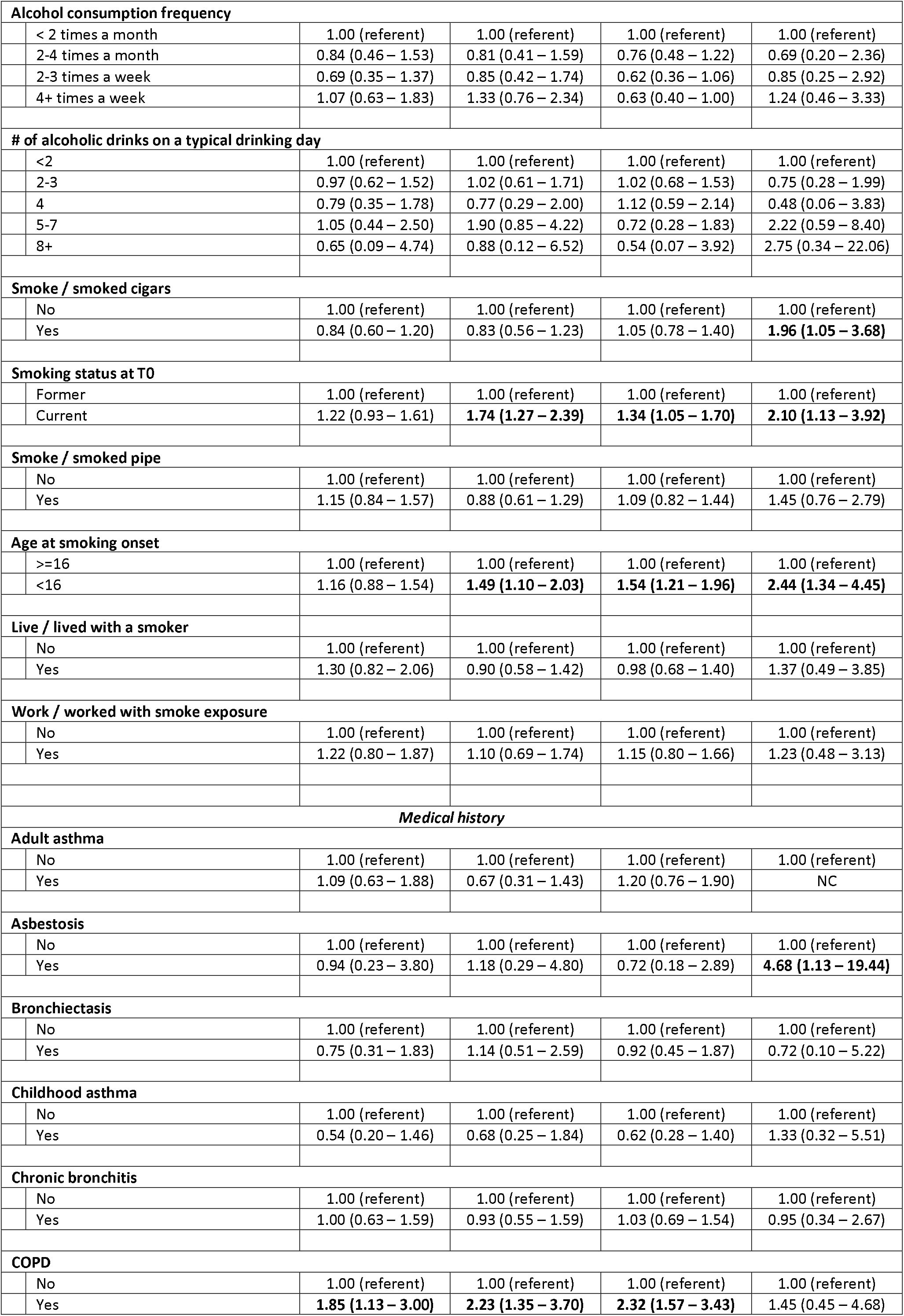
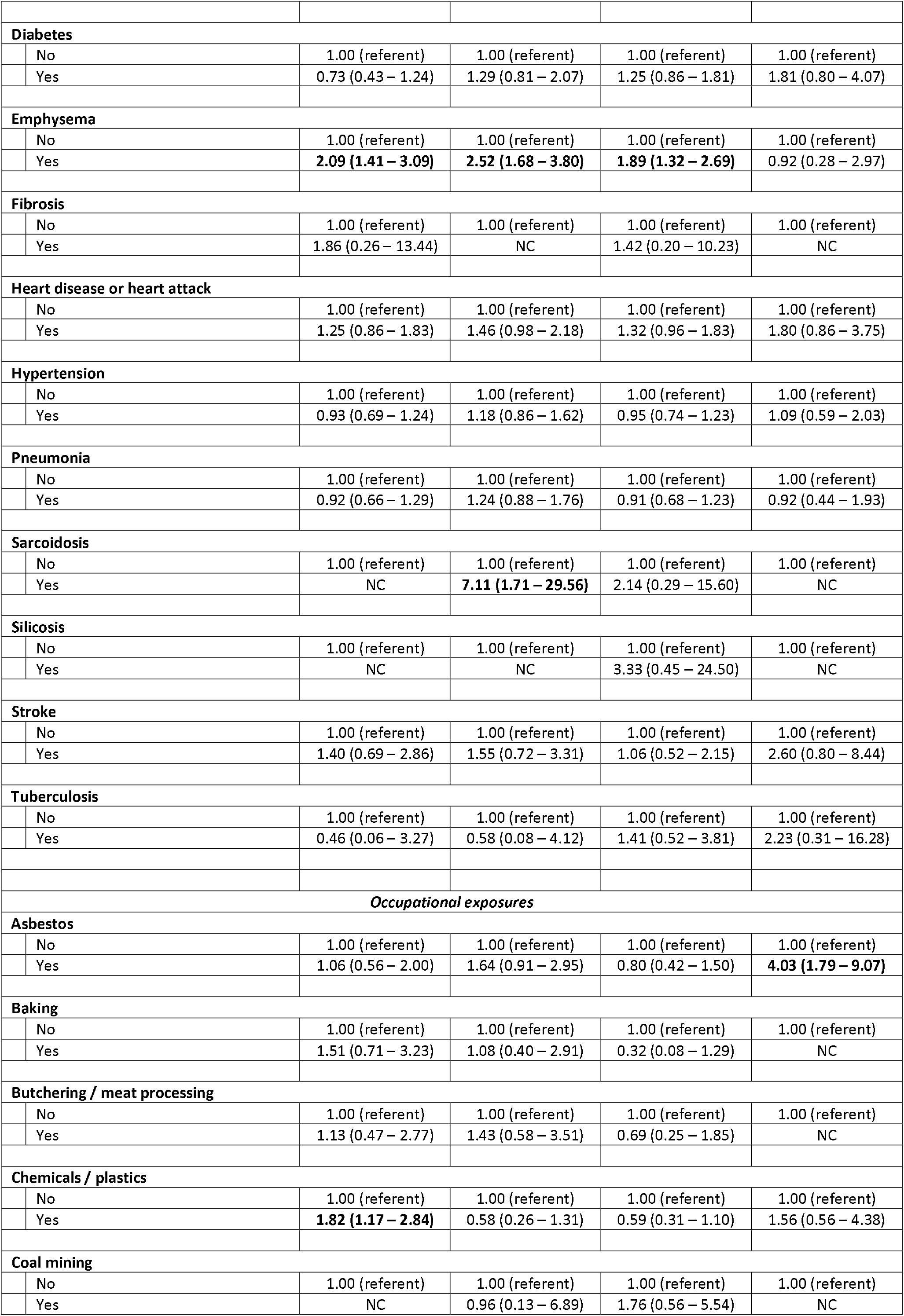
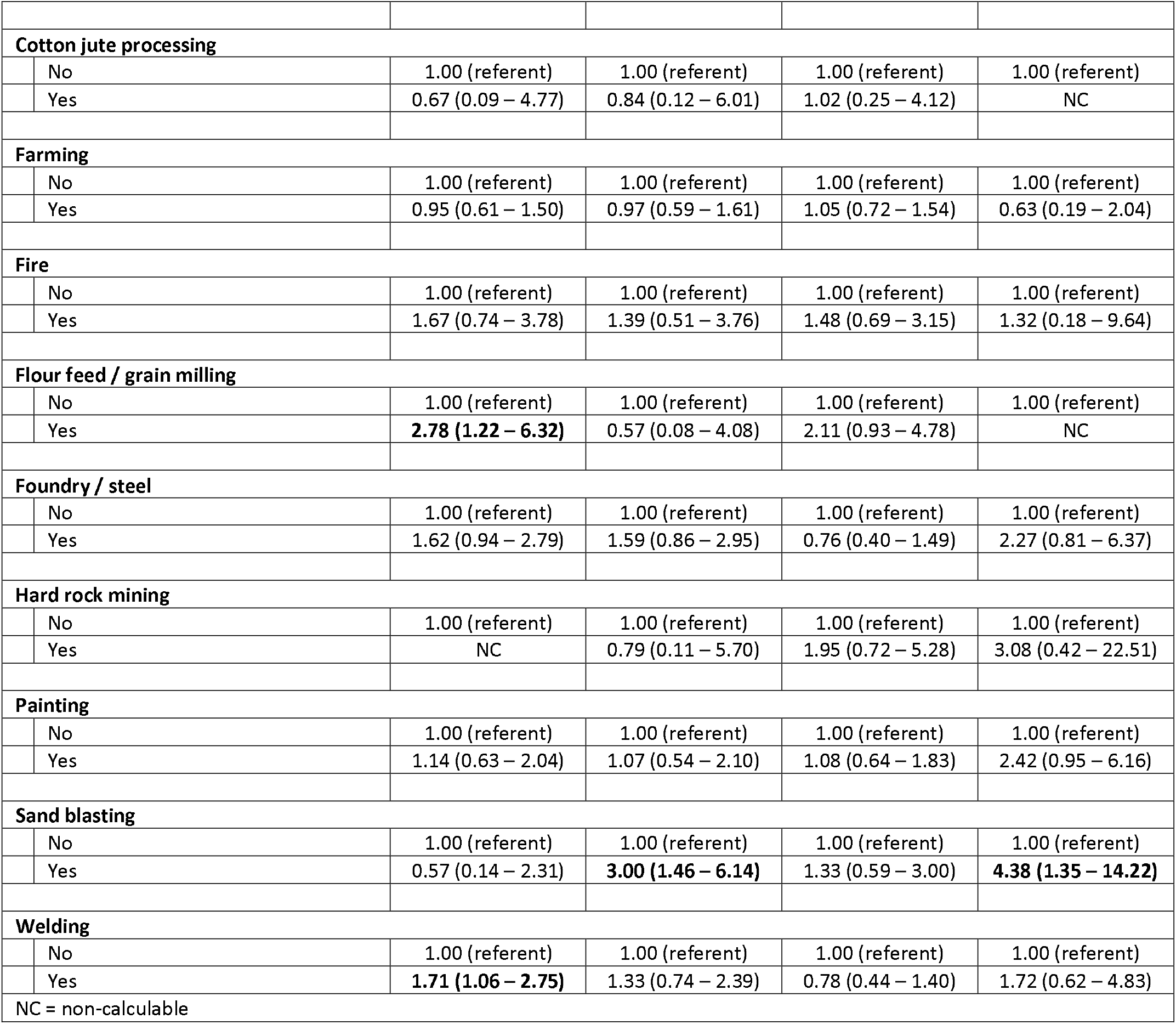
Odds ratios and 95% confidence interval from the 48 univariable logistic regression models.

**Supplemental Table 2.**
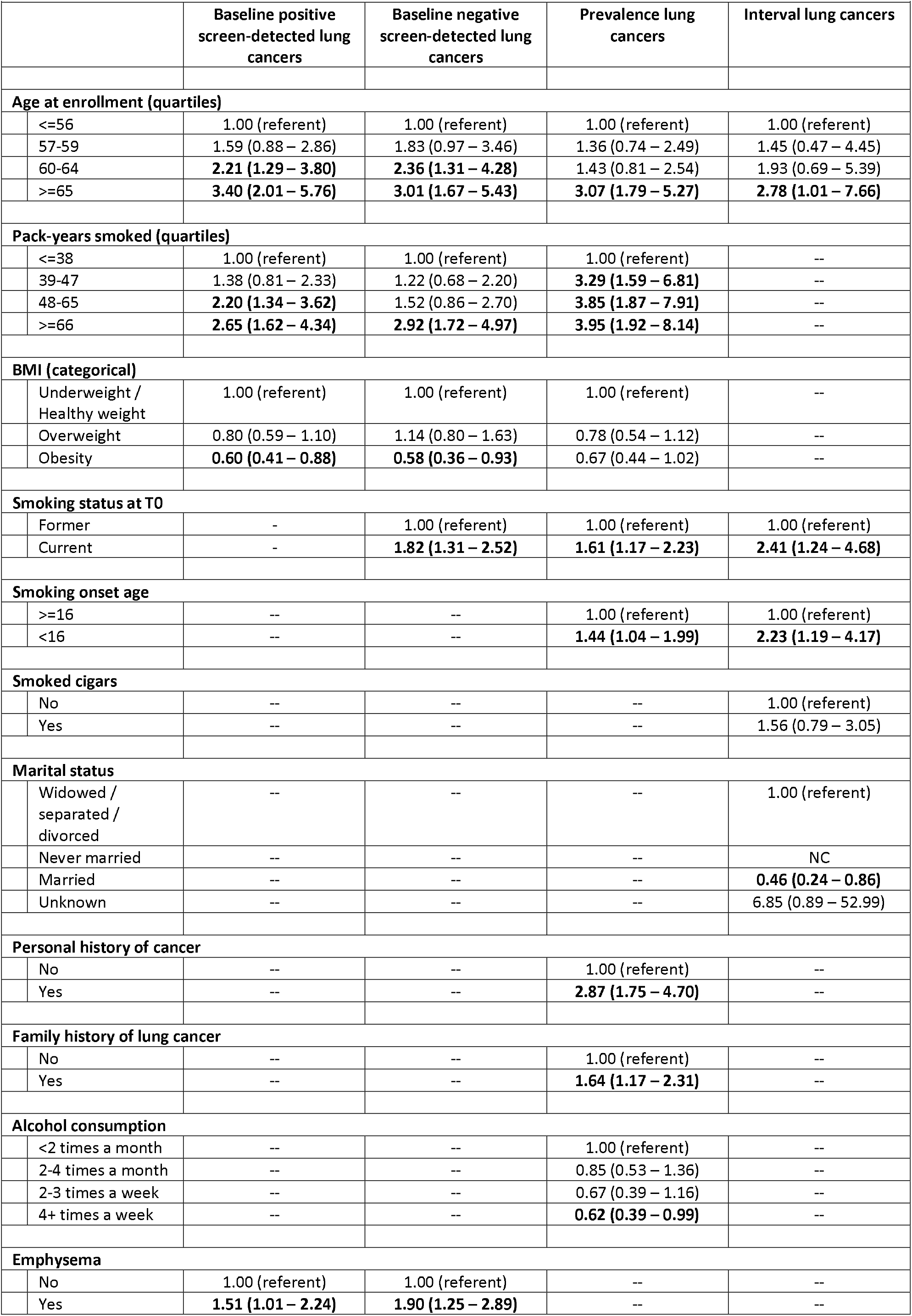
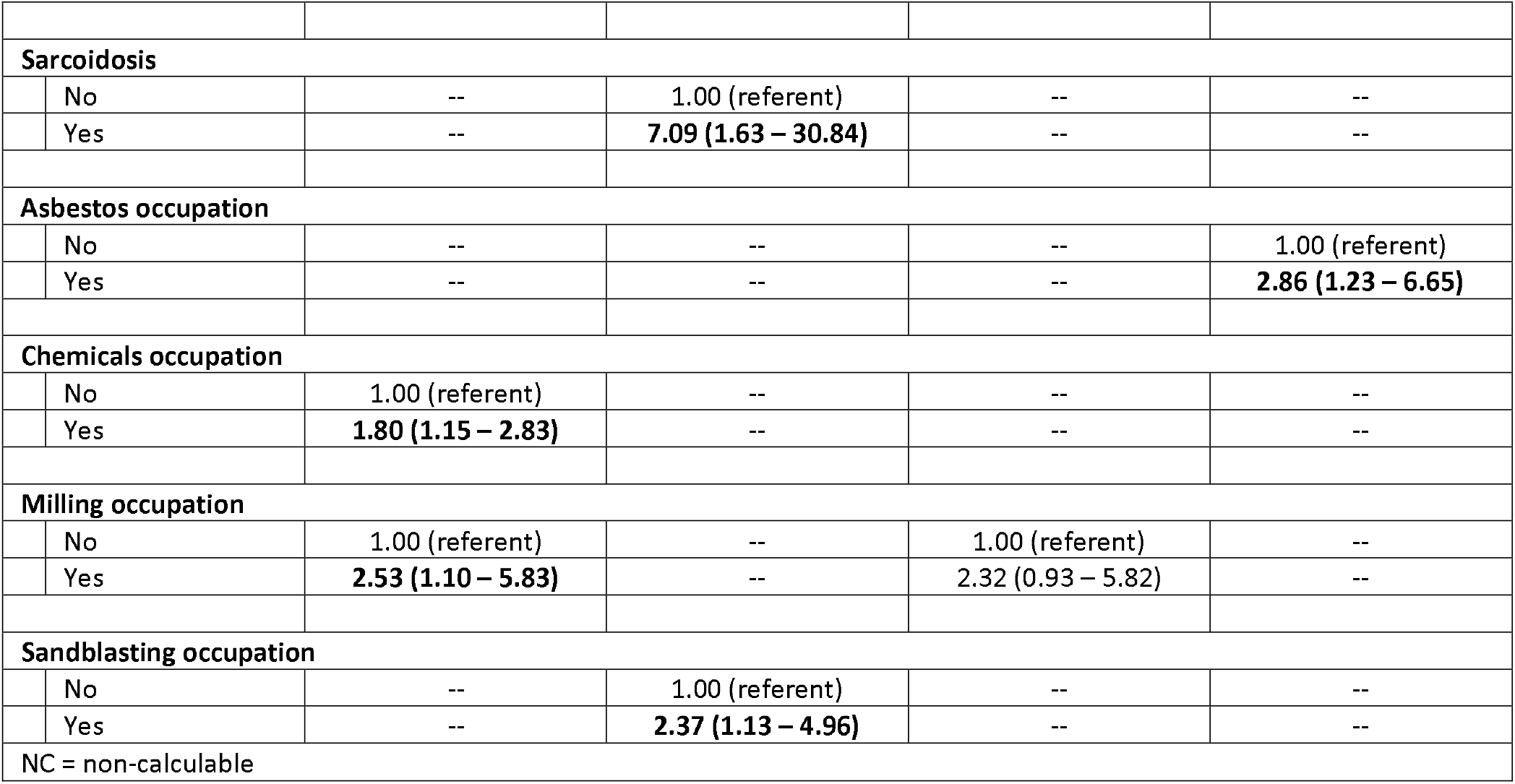
Final risk models adjusted for gender, race, ethnicity, smoke status, and pack-years.

## Notes

### Competing Interest Statement

G.R. reports consulting fees from Impact Business Information Solutions (IBIS) Inc., Princeton, NJ 08542

### Funding Statement

Funding support came from the National Cancer Institute Early Detection Research Network (U01-CA200464 to MBS)

### Author Declarations

This research was approved by the Advarra Institutional Review Board.

