## Supplemental Materials for "Novel risk models based on screening history results and timing of lung cancer diagnosis: *Post hoc* analysis of the National Lung Cancer Screening Trial"

**Results**

Demographic and clinical characteristics of lung cancer cases and controls are presented (**Table 1**). Specifically, controls (mean = 61.4 [SD 5.0]) were statistically significantly younger compared to baseline negative screen-detected lung cancers (mean = 63.4 [SD 5.1]), baseline positive screen-detected lung cancers (mean = 63.7 [SD 5.1]), prevalence lung cancers (mean = 63.8 [SD 5.5]), and interval lung cancers (mean = 63.0 [5.1]). Controls (mean = 55.8 [SD 23.8]) reported a statistically significantly lower number of pack-years smoked compared to baseline negative screen-detected lung cancers (mean = 67.8 [SD 27.9]), baseline positive screen-detected lung cancers (mean = 64.3 [SD 26.8]), prevalence lung cancers (mean = 65.5 [SD 29.8]), and interval lung cancers (mean = 68.0 [27.7]). Controls (mean = 16.7 [SD 3.7]) also reported a statistically significantly higher mean age of smoking onset compared to baseline negative screen-detected lung cancers (mean = 16.1 [SD 3.8]), baseline positive screen-detected lung cancers (mean = 16.2 [SD 3.1]), prevalence lung cancers (mean = 16.2 [SD 3.7]), and interval lung cancers (mean = 15.0 [SD 3.7]). For smoking status, control subjects (47.9%) had a statistically significantly lower percentage of current smokers compared to baseline negative screen-detected lung cancers (61.6%, p-value =0.001), prevalence lung cancers (55.2%, p-value =0.020), and interval lung cancers (65.9%, p-value =0.022). With respect to BMI, controls (mean = 27.9 [SD 5.0]; p-value = 0.002) had significantly higher BMIs on average compared to baseline negative screen-detected lung cancers (mean = 26.6 [SD 4.6]; p-value =0.002), baseline positive screen-detected lung cancers (mean = 26.9 [SD 5.1]; p-value = 0.021), and prevalence lung cancers (mean = 26.8 [SD 4.4]; p-value <0.001). There were no statistically significant differences between controls and the four lung cancer case groups for race and ethnicity.

| **Supplemental Table 1. Odds ratios and 95% confidence interval from the 48 univariable logistic regression models** | | | | | |
| --- | --- | --- | --- | --- | --- |
| **Covariate** | | **Odds ratio (95% confidence interval)** | | | |
| ***Demographics*** | | | | | |
|  | | Baseline positive screen detected lung cancers | Baseline negative screen-detected lung cancers | Prevalence lung cancers | Interval lung cancers |
|  | | OR (95% CI) | OR (95% CI) | OR (95% CI) | OR (95% CI) |
| **Age at enrollment (quartiles)** | | | | | |
|  | <=56 | 1.00 (referent) | 1.00 (referent) | 1.00 (referent) | 1.00 (referent) |
|  | 57-59 | 1.66 (0.93 – 2.99) | **1.90 (1.01 – 3.57)** | 1.29 (0.81 – 2.04) | 1.37 (0.45 – 4.19) |
|  | 60-64 | **2.46 (1.44 – 4.20)** | **2.56 (1.42 – 4.61)** | **1.65 (1.08 – 2.53)** | 1.83 (0.66 – 5.08) |
|  | >=65 | **4.12 (2.45 – 6.93)** | **3.48 (1.95 – 6.22)** | **3.02 (2.02 – 4.52)** | 2.59 (0.96 – 7.03) |
| **Education** | | | | | |
|  | High school or lower | 1.00 (referent) | 1.00 (referent) | 1.00 (referent) | 1.00 (referent) |
|  | Post high school / some college | 0.92 (0.67 – 1.27) | 1.09 (0.76 – 1.57) | **0.74 (0.55 – 0.98)** | 0.66 (0.33 – 1.31) |
|  | Bachelor / grad | 0.71 (0.50 – 1.02) | 0.72 (0.48 – 1.10) | 0.79 (0.59 – 1.06) | 0.47 (0.21 – 1.06) |
|  | Other / N/A | 0.95 (0.38 – 2.37) | 1.03 (0.37 – 2.86) | 0.69 (0.28 – 1.69) | 1.46 (0.34 – 6.31) |
| **Ethnicity** | | | | | |
|  | Neither Hispanic nor Latino | 1.00 (referent) | 1.00 (referent) | 1.00 (referent) | 1.00 (referent) |
|  | Hispanic or Latino (with missing) | 0.20 (0.03 – 1.41) | 0.25 (0.03 – 1.77) | 1.55 (0.82 – 2.93) | 0.94 (0.13 – 6.82) |
|  | Hispanic or Latino (without missing) | NC | 0.33 (0.05 – 2.36) | 1.24 (0.55 – 2.80) | NC |
| **Gender** | | | | | |
|  | Male | 1.00 (referent) | 1.00 (referent) | 1.00 (referent) | 1.00 (referent) |
|  | Female | 1.09 (0.83 – 1.44) | 0.79 (0.57 – 1.08) | 1.03 (0.81 – 1.32) | **0.48 (0.24 – 0.95)** |
| **Race** | | | | | |
|  | White | 1.00 (referent) | 1.00 (referent) | 1.00 (referent) | 1.00 (referent) |
|  | Non-white, NA, or more than one race | 0.56 (0.30 – 1.03) | 1.15 (0.69 – 1.90) | 0.79 (0.50 – 1.26) | 0.99 (0.36 – 2.78) |
| **Pack-years smoked (quartiles)** | | | | | |
|  | <=38 | 1.00 (referent) | 1.00 (referent) | 1.00 (referent) | 1.00 (referent) |
|  | 39-47 | 1.55 (0.92 – 2.62) | 1.51 (0.84 – 2.69) | **3.17 (1.91 – 5.28)** | **5.68 (1.28 – 25.19)** |
|  | 48-65 | **2.64 (1.62 – 4.29)** | **1.89 (1.08 – 3.33)** | **3.88 (2.35 – 6.40)** | 4.60 (1.00 – 21.02) |
|  | >=66 | **3.26 (2.03 – 5.22)** | **3.79 (2.26 – 6.33)** | **4.69 (2.87 – 7.66)** | **8.30 (1.93 – 35.64)** |
| **BMI (categorical)** | | | | | |
|  | Underweight / Healthy weight (<18.5 – 24) | 1.00 (referent) | 1.00 (referent) | 1.00 (referent) | 1.00 (referent) |
|  | Overweight (25-29) | 0.78 (0.57 – 1.07) | 1.07 (0.76 – 1.51) | **0.73 (0.55 – 0.95)** | 0.68 (0.33 – 1.40) |
|  | Obesity (>=30) | **0.59 (0.41 – 0.86)** | **0.52 (0.33 – 0.83)** | **0.52 (0.38 – 0.73)** | 0.93 (0.44 – 1.92) |
| **Marital status** | | | | | |
|  | Widowed/ separated/ divorced | 1.00 (referent) | 1.00 (referent) | 1.00 (referent) | 1.00 (referent) |
|  | Never Married | 0.82 (0.41 – 1.66) | 0.50 (0.18 – 1.38) | 0.98 (0.54 – 1.76) | NC |
|  | Married | 0.85 (0.63 – 1.14) | 0.98 (0.70 – 1.38) | 0.95 (0.73 – 1.24) | **0.51 (0.28 – 0.94)** |
|  | Unknown | 0.85 (0.12 – 6.13) | NC | NC | **5.78 (1.33 – 25.07)** |
| ***Smoking history, Alcohol use, family history of lung cancer, personal history of cancer*** | | | | | |
| **Personal history of any cancer** | | | | | |
|  | No | 1.00 (referent) | 1.00 (referent) | 1.00 (referent) | 1.00 (referent) |
|  | Yes | 1.60 (0.91 – 2.82) | 1.38 (0.70 – 2.71) | **2.43 (1.60 – 3.68)** | 0.57 (0.08 – 4.12) |
| **Family history of lung cancer** | |  |  |  |  |
|  | No | 1.00 (referent) | 1.00 (referent) | 1.00 (referent) | 1.00 (referent) |
|  | Yes | 1.16 (0.84 – 1.60) | 1.09 (0.76 – 1.57) | **1.50 (1.15 – 1.95)** | 0.68 (0.30 – 1.54) |
| **Alcohol consumption frequency** | | | | | |
|  | < 2 times a month | 1.00 (referent) | 1.00 (referent) | 1.00 (referent) | 1.00 (referent) |
|  | 2-4 times a month | 0.84 (0.46 – 1.53) | 0.81 (0.41 – 1.59) | 0.76 (0.48 – 1.22) | 0.69 (0.20 – 2.36) |
|  | 2-3 times a week | 0.69 (0.35 – 1.37) | 0.85 (0.42 – 1.74) | 0.62 (0.36 – 1.06) | 0.85 (0.25 – 2.92) |
|  | 4+ times a week | 1.07 (0.63 – 1.83) | 1.33 (0.76 – 2.34) | 0.63 (0.40 – 1.00) | 1.24 (0.46 – 3.33) |
| **# of alcoholic drinks on a typical drinking day** | | | | | |
|  | <2 | 1.00 (referent) | 1.00 (referent) | 1.00 (referent) | 1.00 (referent) |
|  | 2-3 | 0.97 (0.62 – 1.52) | 1.02 (0.61 – 1.71) | 1.02 (0.68 – 1.53) | 0.75 (0.28 – 1.99) |
|  | 4 | 0.79 (0.35 – 1.78) | 0.77 (0.29 – 2.00) | 1.12 (0.59 – 2.14) | 0.48 (0.06 – 3.83) |
|  | 5-7 | 1.05 (0.44 – 2.50) | 1.90 (0.85 – 4.22) | 0.72 (0.28 – 1.83) | 2.22 (0.59 – 8.40) |
|  | 8+ | 0.65 (0.09 – 4.74) | 0.88 (0.12 – 6.52) | 0.54 (0.07 – 3.92) | 2.75 (0.34 – 22.06) |
| **Smoke / smoked cigars** | | | | | |
|  | No | 1.00 (referent) | 1.00 (referent) | 1.00 (referent) | 1.00 (referent) |
|  | Yes | 0.84 (0.60 – 1.20) | 0.83 (0.56 – 1.23) | 1.05 (0.78 – 1.40) | **1.96 (1.05 – 3.68)** |
| **Smoking status at T0** | | | | | |
|  | Former | 1.00 (referent) | 1.00 (referent) | 1.00 (referent) | 1.00 (referent) |
|  | Current | 1.22 (0.93 – 1.61) | **1.74 (1.27 – 2.39)** | **1.34 (1.05 – 1.70)** | **2.10 (1.13 – 3.92)** |
| **Smoke / smoked pipe** | | | | | |
|  | No | 1.00 (referent) | 1.00 (referent) | 1.00 (referent) | 1.00 (referent) |
|  | Yes | 1.15 (0.84 – 1.57) | 0.88 (0.61 – 1.29) | 1.09 (0.82 – 1.44) | 1.45 (0.76 – 2.79) |
| **Age at smoking onset** | | | | | |
|  | >=16 | 1.00 (referent) | 1.00 (referent) | 1.00 (referent) | 1.00 (referent) |
|  | <16 | 1.16 (0.88 – 1.54) | **1.49 (1.10 – 2.03)** | **1.54 (1.21 – 1.96)** | **2.44 (1.34 – 4.45)** |
| **Live / lived with a smoker** | |  |  |  |  |
|  | No | 1.00 (referent) | 1.00 (referent) | 1.00 (referent) | 1.00 (referent) |
|  | Yes | 1.30 (0.82 – 2.06) | 0.90 (0.58 – 1.42) | 0.98 (0.68 – 1.40) | 1.37 (0.49 – 3.85) |
| **Work / worked with smoke exposure** | | | | | |
|  | No | 1.00 (referent) | 1.00 (referent) | 1.00 (referent) | 1.00 (referent) |
|  | Yes | 1.22 (0.80 – 1.87) | 1.10 (0.69 – 1.74) | 1.15 (0.80 – 1.66) | 1.23 (0.48 – 3.13) |
| ***Medical history*** | | | | | |
| **Adult asthma** | | | | | |
|  | No | 1.00 (referent) | 1.00 (referent) | 1.00 (referent) | 1.00 (referent) |
|  | Yes | 1.09 (0.63 – 1.88) | 0.67 (0.31 – 1.43) | 1.20 (0.76 – 1.90) | NC |
| **Asbestosis** | | | | | |
|  | No | 1.00 (referent) | 1.00 (referent) | 1.00 (referent) | 1.00 (referent) |
|  | Yes | 0.94 (0.23 – 3.80) | 1.18 (0.29 – 4.80) | 0.72 (0.18 – 2.89) | **4.68 (1.13 – 19.44)** |
| **Bronchiectasis** | | | | | |
|  | No | 1.00 (referent) | 1.00 (referent) | 1.00 (referent) | 1.00 (referent) |
|  | Yes | 0.75 (0.31 – 1.83) | 1.14 (0.51 – 2.59) | 0.92 (0.45 – 1.87) | 0.72 (0.10 – 5.22) |
| **Childhood asthma** | | | | | |
|  | No | 1.00 (referent) | 1.00 (referent) | 1.00 (referent) | 1.00 (referent) |
|  | Yes | 0.54 (0.20 – 1.46) | 0.68 (0.25 – 1.84) | 0.62 (0.28 – 1.40) | 1.33 (0.32 – 5.51) |
| **Chronic bronchitis** | | | | | |
|  | No | 1.00 (referent) | 1.00 (referent) | 1.00 (referent) | 1.00 (referent) |
|  | Yes | 1.00 (0.63 – 1.59) | 0.93 (0.55 – 1.59) | 1.03 (0.69 – 1.54) | 0.95 (0.34 – 2.67) |
| **COPD** | | | | | |
|  | No | 1.00 (referent) | 1.00 (referent) | 1.00 (referent) | 1.00 (referent) |
|  | Yes | **1.85 (1.13 – 3.00)** | **2.23 (1.35 – 3.70)** | **2.32 (1.57 – 3.43)** | 1.45 (0.45 – 4.68) |
| **Diabetes** | | | | | |
|  | No | 1.00 (referent) | 1.00 (referent) | 1.00 (referent) | 1.00 (referent) |
|  | Yes | 0.73 (0.43 – 1.24) | 1.29 (0.81 – 2.07) | 1.25 (0.86 – 1.81) | 1.81 (0.80 – 4.07) |
| **Emphysema** | | | | | |
|  | No | 1.00 (referent) | 1.00 (referent) | 1.00 (referent) | 1.00 (referent) |
|  | Yes | **2.09 (1.41 – 3.09)** | **2.52 (1.68 – 3.80)** | **1.89 (1.32 – 2.69)** | 0.92 (0.28 – 2.97) |
| **Fibrosis** | | | | | |
|  | No | 1.00 (referent) | 1.00 (referent) | 1.00 (referent) | 1.00 (referent) |
|  | Yes | 1.86 (0.26 – 13.44) | NC | 1.42 (0.20 – 10.23) | NC |
| **Heart disease or heart attack** | | | | | |
|  | No | 1.00 (referent) | 1.00 (referent) | 1.00 (referent) | 1.00 (referent) |
|  | Yes | 1.25 (0.86 – 1.83) | 1.46 (0.98 – 2.18) | 1.32 (0.96 – 1.83) | 1.80 (0.86 – 3.75) |
| **Hypertension** | | | | | |
|  | No | 1.00 (referent) | 1.00 (referent) | 1.00 (referent) | 1.00 (referent) |
|  | Yes | 0.93 (0.69 – 1.24) | 1.18 (0.86 – 1.62) | 0.95 (0.74 – 1.23) | 1.09 (0.59 – 2.03) |
| **Pneumonia** | | | | | |
|  | No | 1.00 (referent) | 1.00 (referent) | 1.00 (referent) | 1.00 (referent) |
|  | Yes | 0.92 (0.66 – 1.29) | 1.24 (0.88 – 1.76) | 0.91 (0.68 – 1.23) | 0.92 (0.44 – 1.93) |
| **Sarcoidosis** | | | | | |
|  | No | 1.00 (referent) | 1.00 (referent) | 1.00 (referent) | 1.00 (referent) |
|  | Yes | NC | **7.11 (1.71 – 29.56)** | 2.14 (0.29 – 15.60) | NC |
| **Silicosis** | | | | | |
|  | No | 1.00 (referent) | 1.00 (referent) | 1.00 (referent) | 1.00 (referent) |
|  | Yes | NC | NC | 3.33 (0.45 – 24.50) | NC |
| **Stroke** | | | | | |
|  | No | 1.00 (referent) | 1.00 (referent) | 1.00 (referent) | 1.00 (referent) |
|  | Yes | 1.40 (0.69 – 2.86) | 1.55 (0.72 – 3.31) | 1.06 (0.52 – 2.15) | 2.60 (0.80 – 8.44) |
| **Tuberculosis** | | | | | |
|  | No | 1.00 (referent) | 1.00 (referent) | 1.00 (referent) | 1.00 (referent) |
|  | Yes | 0.46 (0.06 – 3.27) | 0.58 (0.08 – 4.12) | 1.41 (0.52 – 3.81) | 2.23 (0.31 – 16.28) |
| ***Occupational exposures*** | | | | | |
| **Asbestos** | | | | | |
|  | No | 1.00 (referent) | 1.00 (referent) | 1.00 (referent) | 1.00 (referent) |
|  | Yes | 1.06 (0.56 – 2.00) | 1.64 (0.91 – 2.95) | 0.80 (0.42 – 1.50) | **4.03 (1.79 – 9.07)** |
| **Baking** | | | | | |
|  | No | 1.00 (referent) | 1.00 (referent) | 1.00 (referent) | 1.00 (referent) |
|  | Yes | 1.51 (0.71 – 3.23) | 1.08 (0.40 – 2.91) | 0.32 (0.08 – 1.29) | NC |
| **Butchering / meat processing** | | | | | |
|  | No | 1.00 (referent) | 1.00 (referent) | 1.00 (referent) | 1.00 (referent) |
|  | Yes | 1.13 (0.47 – 2.77) | 1.43 (0.58 – 3.51) | 0.69 (0.25 – 1.85) | NC |
| **Chemicals / plastics** | | | | | |
|  | No | 1.00 (referent) | 1.00 (referent) | 1.00 (referent) | 1.00 (referent) |
|  | Yes | **1.82 (1.17 – 2.84)** | 0.58 (0.26 – 1.31) | 0.59 (0.31 – 1.10) | 1.56 (0.56 – 4.38) |
| **Coal mining** | | | | | |
|  | No | 1.00 (referent) | 1.00 (referent) | 1.00 (referent) | 1.00 (referent) |
|  | Yes | NC | 0.96 (0.13 – 6.89) | 1.76 (0.56 – 5.54) | NC |
| **Cotton jute processing** | | | | | |
|  | No | 1.00 (referent) | 1.00 (referent) | 1.00 (referent) | 1.00 (referent) |
|  | Yes | 0.67 (0.09 – 4.77) | 0.84 (0.12 – 6.01) | 1.02 (0.25 – 4.12) | NC |
| **Farming** | | | | | |
|  | No | 1.00 (referent) | 1.00 (referent) | 1.00 (referent) | 1.00 (referent) |
|  | Yes | 0.95 (0.61 – 1.50) | 0.97 (0.59 – 1.61) | 1.05 (0.72 – 1.54) | 0.63 (0.19 – 2.04) |
| **Fire** | | | | | |
|  | No | 1.00 (referent) | 1.00 (referent) | 1.00 (referent) | 1.00 (referent) |
|  | Yes | 1.67 (0.74 – 3.78) | 1.39 (0.51 – 3.76) | 1.48 (0.69 – 3.15) | 1.32 (0.18 – 9.64) |
| **Flour feed / grain milling** | | | | | |
|  | No | 1.00 (referent) | 1.00 (referent) | 1.00 (referent) | 1.00 (referent) |
|  | Yes | **2.78 (1.22 – 6.32)** | 0.57 (0.08 – 4.08) | 2.11 (0.93 – 4.78) | NC |
| **Foundry / steel** | | | | | |
|  | No | 1.00 (referent) | 1.00 (referent) | 1.00 (referent) | 1.00 (referent) |
|  | Yes | 1.62 (0.94 – 2.79) | 1.59 (0.86 – 2.95) | 0.76 (0.40 – 1.49) | 2.27 (0.81 – 6.37) |
| **Hard rock mining** | | | | | |
|  | No | 1.00 (referent) | 1.00 (referent) | 1.00 (referent) | 1.00 (referent) |
|  | Yes | NC | 0.79 (0.11 – 5.70) | 1.95 (0.72 – 5.28) | 3.08 (0.42 – 22.51) |
| **Painting** | | | | | |
|  | No | 1.00 (referent) | 1.00 (referent) | 1.00 (referent) | 1.00 (referent) |
|  | Yes | 1.14 (0.63 – 2.04) | 1.07 (0.54 – 2.10) | 1.08 (0.64 – 1.83) | 2.42 (0.95 – 6.16) |
| **Sand blasting** | | | | | |
|  | No | 1.00 (referent) | 1.00 (referent) | 1.00 (referent) | 1.00 (referent) |
|  | Yes | 0.57 (0.14 – 2.31) | **3.00 (1.46 – 6.14)** | 1.33 (0.59 – 3.00) | **4.38 (1.35 – 14.22)** |
| **Welding** | | | | | |
|  | No | 1.00 (referent) | 1.00 (referent) | 1.00 (referent) | 1.00 (referent) |
|  | Yes | **1.71 (1.06 – 2.75)** | 1.33 (0.74 – 2.39) | 0.78 (0.44 – 1.40) | 1.72 (0.62 – 4.83) |
| NC = non-calculable | | | | | |

| **Supplemental Table 2. Final risk models adjusted for gender, race, ethnicity, smoke status, and pack-years** | | | | | |
| --- | --- | --- | --- | --- | --- |
|  | | **Baseline positive screen-detected lung cancers** | **Baseline negative screen-detected lung cancers** | **Prevalence lung cancers** | **Interval lung cancers** |
| **Age at enrollment (quartiles)** | | | | | |
|  | <=56 | 1.00 (referent) | 1.00 (referent) | 1.00 (referent) | 1.00 (referent) |
|  | 57-59 | 1.59 (0.88 – 2.86) | 1.83 (0.97 – 3.46) | 1.36 (0.74 – 2.49) | 1.45 (0.47 – 4.45) |
|  | 60-64 | **2.21 (1.29 – 3.80)** | **2.36 (1.31 – 4.28)** | 1.43 (0.81 – 2.54) | 1.93 (0.69 – 5.39) |
|  | >=65 | **3.40 (2.01 – 5.76)** | **3.01 (1.67 – 5.43)** | **3.07 (1.79 – 5.27)** | **2.78 (1.01 – 7.66)** |
| **Pack-years smoked (quartiles)** | | | | | |
|  | <=38 | 1.00 (referent) | 1.00 (referent) | 1.00 (referent) | -- |
|  | 39-47 | 1.38 (0.81 – 2.33) | 1.22 (0.68 – 2.20) | **3.29 (1.59 – 6.81)** | -- |
|  | 48-65 | **2.20 (1.34 – 3.62)** | 1.52 (0.86 – 2.70) | **3.85 (1.87 – 7.91)** | -- |
|  | >=66 | **2.65 (1.62 – 4.34)** | **2.92 (1.72 – 4.97)** | **3.95 (1.92 – 8.14)** | -- |
| **BMI (categorical)** | | | | | |
|  | Underweight / Healthy weight | 1.00 (referent) | 1.00 (referent) | 1.00 (referent) | -- |
|  | Overweight | 0.80 (0.59 – 1.10) | 1.14 (0.80 – 1.63) | 0.78 (0.54 – 1.12) | -- |
|  | Obesity | **0.60 (0.41 – 0.88)** | **0.58 (0.36 – 0.93)** | 0.67 (0.44 – 1.02) | -- |
| **Smoking status at T0** | | | | | |
|  | Former | - | 1.00 (referent) | 1.00 (referent) | 1.00 (referent) |
|  | Current | - | **1.82 (1.31 – 2.52)** | **1.61 (1.17 – 2.23)** | **2.41 (1.24 – 4.68)** |
| **Smoking onset age** | | | | | |
|  | >=16 | -- | -- | 1.00 (referent) | 1.00 (referent) |
|  | <16 | -- | -- | **1.44 (1.04 – 1.99)** | **2.23 (1.19 – 4.17)** |
| **Smoked cigars** | | | | | |
|  | No | -- | -- | -- | 1.00 (referent) |
|  | Yes | -- | -- | -- | 1.56 (0.79 – 3.05) |
| **Marital status** | | | | | |
|  | Widowed / separated / divorced | -- | -- | -- | 1.00 (referent) |
|  | Never married | -- | -- | -- | NC |
|  | Married | -- | -- | -- | **0.46 (0.24 – 0.86)** |
|  | Unknown | -- | -- | -- | 6.85 (0.89 – 52.99) |
| **Personal history of cancer** | | | | | |
|  | No | -- | -- | 1.00 (referent) | -- |
|  | Yes | -- | -- | **2.87 (1.75 – 4.70)** | -- |
| **Family history of lung cancer** | | | | | |
|  | No | -- | -- | 1.00 (referent) | -- |
|  | Yes | -- | -- | **1.64 (1.17 – 2.31)** | -- |
| **Alcohol consumption** | | | | | |
|  | <2 times a month | -- | -- | 1.00 (referent) | -- |
|  | 2-4 times a month | -- | -- | 0.85 (0.53 – 1.36) | -- |
|  | 2-3 times a week | -- | -- | 0.67 (0.39 – 1.16) | -- |
|  | 4+ times a week | -- | -- | **0.62 (0.39 – 0.99)** | -- |
| **Emphysema** | | | | | |
|  | No | 1.00 (referent) | 1.00 (referent) | -- | -- |
|  | Yes | **1.51 (1.01 – 2.24)** | **1.90 (1.25 – 2.89)** | -- | -- |
| **Sarcoidosis** | | | | | |
|  | No | -- | 1.00 (referent) | -- | -- |
|  | Yes | -- | **7.09 (1.63 – 30.84)** | -- | -- |
| **Asbestos occupation** | | | | | |
|  | No | -- | -- | -- | 1.00 (referent) |
|  | Yes | -- | -- | -- | **2.86 (1.23 – 6.65)** |
| **Chemicals occupation** | | | | | |
|  | No | 1.00 (referent) | -- | -- | -- |
|  | Yes | **1.80 (1.15 – 2.83)** | -- | -- | -- |
| **Milling occupation** | | | | | |
|  | No | 1.00 (referent) | -- | 1.00 (referent) | -- |
|  | Yes | **2.53 (1.10 – 5.83)** | -- | 2.32 (0.93 – 5.82) | -- |
| **Sandblasting occupation** | | | | | |
|  | No | -- | 1.00 (referent) | -- | -- |
|  | Yes | -- | **2.37 (1.13 – 4.96)** | -- | -- |
| NC = non-calculable | | | | | |
